# Efficacy of Corticosteroids in Non-Intensive Care Unit Patients with COVID-19 Pneumonia from the New York Metropolitan region

**DOI:** 10.1101/2020.07.02.20145565

**Authors:** Monil Majmundar, Tikal Kansara, Joanna Marta Lenik, Hansang Park, Kuldeep Ghosh, Rajkumar Doshi, Palak Shah, Ashish Kumar, Hossam Amin, Shobhana Chaudhari, Imnett Habtes

## Abstract

**Introduction:** The role of systemic corticosteroid as a therapeutic agent for patients with COVID-19 pneumonia is controversial.

**Objective:** The purpose of this study was to evaluate the effect of corticosteroids in non-intensive care unit (ICU) patients with COVID-19 pneumonia complicated by acute hypoxemic respiratory failure (AHRF).

**Methods:** This was a single-center retrospective cohort study, from the 16^th^ March, 2020 to 30^th^ April, 2020; final follow-up on 10^th^ May, 2020. 265 patients consecutively admitted to the non-ICU wards with laboratory-confirmed COVID-19 pneumonia were screened for inclusion. 205 patients who developed AHRF (SpO_2_/FiO_2_ ≤ 440 or PaO_2_/FiO_2_ ≤ 300) were only included in the final study. Direct admission to the Intensive care unit (ICU), patients developing composite primary outcome within 24 hours of admission, and patients who never became hypoxic during their stay in the hospital were excluded. Patients divided into two cohort based on corticosteroid. The primary outcome was a composite of ICU transfer, intubation, or in-hospital mortality. Secondary outcomes were ICU transfer, intubation, in-hospital mortality, discharge, length of stay and daily trend of SpO_2_/FiO_2_ (SF) ratio from the index date. Cox-proportional hazard regression was implemented to analyze the time to event outcomes.

**Result:** Among 205 patients, 60 (29.27%) were treated with corticosteroid. The mean age was ∼57 years, and ∼75% were men. Thirteen patients (22.41%) developed a primary composite outcome in the corticosteroid cohort vs. 54 (37.5%) patients in the non-corticosteroid cohort (P=0.039). The adjusted hazard ratio (HR) for the development of the composite primary outcome was 0.15 (95% CI, 0.07 – 0.33; P <0.001). The adjusted hazard ratio for ICU transfer was 0.16 (95% CI, 0.07 to 0.34; P < 0.001), intubation was 0.31 (95% CI, 0.14 to 0.70; P – 0.005), death was 0.53 (95% CI, 0.22 to 1.31; P – 0.172), and discharge was 3.65 (95% CI, 2.20 to 6.06; P<0.001). The corticosteroid cohort had increasing SpO_2_/FiO_2_ over time compared to the non-corticosteroid cohort who experience decreasing SpO_2_/FiO_2_ over time.

**Conclusion:** Among non-ICU patients hospitalized with COVID-19 pneumonia complicated by AHRF, treatment with corticosteroid was associated with a significantly lower risk of the primary composite outcome of ICU transfer, intubation, or in-hospital death.

## Introduction

Severe Acute Respiratory Syndrome Coronavirus 2 (SARS-CoV-2) is the RNA virus that causes coronavirus disease 2019 (COVID-19). This virus is responsible for a spectrum of disease presentation, which ranges from asymptomatic infection to severe pneumonia, respiratory failure, and death.^1^ To date, SARS-CoV-2 has caused a significant degree of morbidity and mortality within the United States, with a large proportion of these cases concentrated in New York City.^1^ Given the novelty of this virus, there is limited data on treating these patients, and at the current time, there is no definitive treatment for COVID-19 infection. As a result, management protocols vary and are rapidly evolving with emerging data and clinical experiences.

The use of systemic corticosteroids in the management of COVID-19 infection is widely debated. The use of corticosteroids with influenza pneumonia has previously been associated with a higher risk of death ^2,3^ and delayed viral clearance during Severe Acute Respiratory Syndrome-related coronavirus (SARS) and Middle East Respiratory Syndrome (MERS) outbreaks. ^4–6^ Alternatively, few studies supported the use of corticosteroids at a low-to-moderate dose in patients with coronavirus and Influenza A (H1N1) and SARS pneumonia. ^7,8^ On the other hand, the World Health Organization has recommended against routine use of systemic corticosteroids to patients with COVID-19 ^9^; nevertheless, a consensus statement by the Chinese Thoracic Society recommends judicious use of corticosteroids in these patients.^10^

To address this conundrum, we have performed a retrospective cohort study on patients admitted to the general inpatient wards with a diagnosis of COVID-19 pneumonia complicated by acute hypoxemic respiratory failure. The purpose of this study was to determine the clinical efficacy of corticosteroids on outcomes of intensive care unit (ICU) transfer, intubation, in-hospital death, discharge, and length of stay. We hypothesized that systemic corticosteroid use would be associated with a lower risk of a composite endpoint of ICU transfer, intubation, or death.

## Methods

### Study Population

This is a retrospective cohort study of confirmed cases of COVID-19 pneumonia hospitalized at Metropolitan Hospital Center serving the East Harlem community in New York City. All patients were diagnosed with COVID-19 pneumonia as per the World Health Organization’s interim guidance document.^11^ We collected information on consecutive patients admitted to the general wards in Metropolitan Hospital from March 15, 2020, to April 30, 2020, as per our inclusion and exclusion criteria. The ethics committee of New York City Health and Hospital (STAR) and BRANY institution review board approved this study and permitted a waiver of informed consent from the study participant.

Patients were eligible for the study if they met the following inclusion criteria are 1) Age ≥ 18 years old, 2) Confirmed cases of SARS-CoV-2 by PCR method, 3) Admitted in general wards, 4) PaO2/FiO2 (PF) ratio <300 if Arterial blood gas if available or SpO_2_/Fio_2_ (SF) ratio <440, 5) Bilateral infiltrate on chest imaging by radiology staff. The exclusion criteria are 1) Patients with severe immunosuppression (HIV infection, long term use of immunosuppressive agents), 2) Pregnant woman or Lactation period, 3) Oral glucocorticoids were needed for other diseases, 4) Direct admission to intensive care unit (ICU), 5) if had any of primary composite outcome within first 24 hours of admission, 6) Patient who never required oxygen during the hospital course, 7) Patients who left against medical advice.

### Procedure

A team of resident physicians reviewed and collected demographic, laboratory, clinical, and outcomes data from electronic medical records between March 15 and April 30, 2020. The inclusion criteria, exclusion criteria, individual components of all definitions of clinical outcomes were recorded separately and checked by two authors (M.M. and I.H.) (**Supplemental file Appendix 1**). Two independent residents adjudicated all the outcome data, and any disparity was resolved by consulting the primary investigator. Patient confidentiality was protected by allocating a deidentified patient identification, and the electronic data was stored in a locked, password-protected computer.

Nasopharyngeal swab samples were obtained from all patients at admission and tested using real-time reverse transcriptase-polymerase chain reaction assays at LabCorp laboratory to identify SARS-CoV-2 infected patients. The decision to give corticosteroids was at the discretion of the treating physician. All corticosteroid dosages were converted to the equivalent dose of methylprednisolone.^12^ In the non-corticosteroid cohort, the index date was taken as the date when the patient’s SF ratio went below 440, or the PF ratio went below 300. For the corticosteroid cohort, the date when corticosteroid was started was taken as the index date. The calculation of SF ratio and PF ratio is mentioned in the **supplementary file Appendix 2**.

### Outcomes

Our primary outcome was the composite outcome of intensive care unit (ICU) transfer, intubation, or death. Secondary outcomes were discharge, intensive care unit transfer, intubation, death, length of stay, and a daily trend of SF ratio since the index date.

### Statistical Analysis

Baseline characteristics of both cohorts were expressed using descriptive statistics. The continuous variables were exhibited as a mean ± standard deviation or a median with interquartile range (IQR) for normal and non-normal distribution, respectively. Categorical variables were extrapolated in frequency and proportions. The t-test and Mann-Whitney-Wilcoxon tests were applied for normal and non-normal distribution, respectively, to compare continuous variables between two cohorts. Fisher’s exact test or Pearson’s chi^2^ tests were implemented to compare categorical variables.

Time to event (composite primary outcome and secondary outcomes) was defined as the time from the index date of the study to the specified events. The index date was defined as the date when corticosteroid was commenced in the corticosteroid cohort and the day when patients’ inclusion criteria were met in the non-corticosteroid cohort to minimize the difference in the severity between two cohorts. We used the Cox-proportional hazard model to determine univariable and multivariable hazard ratio (HR) and 95% confidence interval (CI) for the corticosteroid group compared to the non-corticosteroid group on the development of composite primary outcome, ICU transfer, intubation, death, and discharge. The multivariable Cox-proportional model included SF ratio, age, gender, chronic lung disease, white blood cell count, platelet count, tocilizumab, and therapeutic dose of Enoxaparin (for the construction of multivariate model - **Supplementary file Appendix 3**). We applied the test for proportionality assumption based on the Schoenfeld Residuals. For the primary outcome and other secondary outcomes, we constructed Kaplan-Meier curves and used the log-rank test to compare. Censoring was applied on the day of discharge. Linear regression was used to show the difference between the length of stay between two cohorts. Mixed-effects linear regression with restricted maximum likelihood was used to perform a longitudinal analysis to assess the trend of SF ratio over time between two cohorts. The fixed-effect constant was suppressed, and unstructured residual errors were generated. A likelihood ratio test vs. linear model chi-square test was used to assess the appropriateness of using a multilevel model. Margins were calculated to display predicted probabilities. Missing data were not imputed. All tests were 2-sided, and a P-value less than 0.05 was considered statistically significant. All analyses were performed with STATA software, version 16.0 (StataCorp LLC).

## Result

### Study population

We included 205 consecutive patients admitted to the Metropolitan Hospital, New York Medical College, from March 15 to April 30, 2020, in this study. The date of the final follow-up was May 10, 2020. The mean age of the entire cohort was 57.61±15.86 years, 153 (74.63%) patients were male, and 149 (73.04%) patients were of Hispanic ethnicity/race. The common comorbidities were hypertension (103, 50.24%), obesity (84, 42%), and diabetes (83, 40.49%) **(Table 1)**. At the time of analysis, a total of 14 patients (out of 205) were still admitted to the hospital. Of 205 patients, 60 (29.27%) patients received systemic corticosteroids, and 145 (70.73%) patients did not receive it.

**Table 1.** Demographic, laboratory characteristics, and outcomes by Corticosteroids. **Abbreviations:** BMI – Basal Metabolic Index, Hb – Hemoglobin, WBC – white blood cell, PaO2 – Partial Pressure of Arterial Oxygen, SpO2 = Saturation of oxygen from Pulse oximeter, AST – Aspartate Transferase, ALT – Alanine Transferase, LDH – Lactate Dehydrogenase, IL-6 – Interleukin-6, ICU – Intensive Care Unit, SOFA – Sequential organ failure assessment, IQR – Interquartile range $ - calculated by converting SpO_2_ to PaO_2_, then diving by FiO_2_ # - includes Hydrocortisone, Prednisone, Methylprednisolone, Dexamethasone

|  | Total<br>(N=205) | Corticosteroids<br>(N=60) | Without corticosteroids<br>(N=145) | P-value |
| --- | --- | --- | --- | --- |
| Demographic Characteristics |  |  |  |  |
| Age, mean (SD) | 57.61 (15.86) | 58.67 (13.35) | 57.18 (16.81) | 0.543 |
| Sex, n (%) |  |  |  | 0.011 |
| Male | 153 (74.63) | 52 (86.67) | 101 (69.67) |  |
| Female | 52 (25.37) | 8 (13.33) | 44 (30.34) |  |
| Ethnicity/Race, n (%) |  |  |  | 0.476 |
| Hispanic | 149 (73.04) | 46 (77.97) | 103 (71.03) |  |
| Non-Hispanic African American | 29 (14.22) | 5 (8.47) | 24 (16.55) |  |
| Non-Hispanic White | 8 (3.92) | 3 (5.08) | 5 (3.45) |  |
| Others | 18 (8.82) | 5 (8.47) | 13 (8.97) |  |
| Comorbidities |  |  |  |  |
| Obesity, n (%) | 84 (42) | 18/59 (30.51) | 66/141 (46.81) | 0.033 |
| BMI (kg/m<sup>2</sup>), median (IQR) | 28.7 (25-33.47) | 26.7 (23.1-31.8) | 29.34 (25-34.64) | 0.035 |
| Hypertension, n (%) | 103 (50.24) | 35 (58.33) | 68 (46.90) | 0.136 |
| Diabetes Mellitus, n (%) | 83 (40.49) | 23 (38.33) | 60 (41.38) | 0.686 |
| Chronic Lung disease, n (%) | 32 (15.61) | 11 (18.33) | 21 (14.48) | 0.489 |
| Cancer, n (%) | 10 (4.88) | 1 (1.67) | 9 (6.21) | 0.170 |
| Coronary Disease, n (%) | 23 (11.22) | 5 (8.33) | 18 (12.41) | 0.40 |
| Heart Failure, n (%) | 7 (3.41) | 2 (3.33) | 5 (3.45) | 0.967 |
| Clinical Parameters |  |  |  |  |
| PaO2/FiO2, median (IQR)<sup>§</sup> | 239.29<br>(152-277.78) | 136.36<br>(65.13-218.97) | 261.91<br>(219.04-280.95) | <0.001 |
| SpO2/FiO2, median (IQR) | 328.57<br>(213.64-409.52)) | 190<br>(92.5-298.44) | 339.29<br>(278.13-419.05) | <0.001 |
| SOFA score < 5, n (%) |  | 53 (88.33) | 120 (82.76) | 0.40 |
| SOFA score ≥ 5, n (%) |  | 7 (11.67) | 25 (17.24) |  |
| Medications |  |  |  |  |
| Corticosteroid<sup>#</sup>, median (IQR) |  |  |  |  |
| Initiation from admission<br>(Days) |  | 2 (1-5) | - | - |
| Dose (mg/day) |  | 80 (60-107) | - |  |
| Duration (Days) |  | 5 (4-7) | - |  |
| Total Dose (mg) |  | 400 (240-642) | - |  |
| Hydroxychloroquine, n (%) | 201 (98.05) | 57 (95) | 144 (99.31) | **0.076** |
| Tocilizumab, n (%) | 18 (8.78) | 11 (18.33) | 7 (4.83) | **0.002** |
| Enoxaparin therapeutic dose, n (%) | 76 (37.07) | 40 (66.67) | 36 (24.83) | **<0.001** |
| **Laboratory Parameters** |  |  |  |  |
| Hb (g/dL), mean (SD) | 12.84 (2.10) | 13 (1.77) | 12.78 (2.23) | 0.492 |
| WBC ($\times 10^3/\mu\text{L}$), median (IQR) | 7.92 (5.97-10.8) | 8.61 (6.42-12.03) | 7.68 (5.8-10.48) | 0.107 |
| Absolute Lymphocyte Count ($\times 10^3/\mu\text{L}$), median (IQR) | 0.89 (0.64-1.25) | 0.84 (0.63-1.24) | 0.92 (0.65-1.31) | 0.427 |
| Platelets ($\times 10^9/\text{L}$), median (IQR) | 211 (163-301) | 278.5 (185-338.50) | 200 (157-260) | **<0.001** |
| GFR (ml/min/1.73m<sup>2</sup>), median (IQR) | 77.3 (50-112.5) | 70.10 (60-105.60) | 79 (41-113.90) | 0.939 |
| AST (units/L), median (IQR) | 46.50 (34-65) | 55 (36-72) | 44 (33-61) | **0.048** |
| ALT (units/L), median (IQR) | 33 (21-64) | 42 (23-95) | 32 (21-54) | 0.058 |
| **Inflammatory Markers** |  |  |  |  |
| Procalcitonin (ng/mL), median (IQR), (N=160) | 0.33 (0.17-0.73) | 0.42 (0.17-0.89) | 0.29 (0.16-0.60) | 0.301 |
| Ferritin (ng/mL), median (IQR), (N=150) | 987 (529-1685) | 1002 (652-1774) | 982 (457-1631) | 0.291 |
| LDH (U/L), median (IQR), (N=148) | 404 (300.47-485.5) | 449 (325-540) | 386 (297-475) | 0.068 |
| D-Dimer (ng/dL), median (IQR), (N=164) | 443 (261-1196) | 653.5 (347.5-2065.5) | 377 (235.50-1015) | **0.004** |
| C-Reactive Protein (mg/dL), median (IQR), (N=135) | 16.24 (7.36-24.38) | 21 (9.76-29.34) | 12.1 (6.65-22.19) | **0.005** |
| IL-6 (pg/ml), median (IQR), (N=36) | 87.25 (28.90-146.25) | 65 (25.80-104) | 116 (89.30-182.10) | 0.084 |
| **Outcomes, n (%)** |  |  |  |  |
| ICU transfer/Intubation /Death, (N=202) | 67 (33.17) | 13 (22.41) | 54 (37.50) | **0.039** |
| ICU Transfer, (N=202) | 59 (29.21) | 12 (20.69) | 47 (32.64) | 0.091 |
| Intubation, (N=200) | 47 (23.50) | 11 (18.97) | 36 (25.35) | 0.334 |
| Death, (N=191) | 42 (21.99) | 8 (14.55) | 34 (25) | 0.114 |
| Discharge, (N=191) | 149 (78.01) | 47 (85.45) | 102 (75) | 0.114 |
| Length of stay (days), median (IQR), (N=191) | 8 (5-15) | 9 (6-17) | 7 (5-13.50) | **0.025** |

### Corticosteroid cohort

Patients in the corticosteroid cohort received systemic corticosteroids in the form of methylprednisolone (n=29, 48.33%), prednisone (n=10, 16.67%), hydrocortisone (n=1, 1.67%), and dexamethasone (n=20, 33.33%). Corticosteroid was commenced at a median of 2 days (IQR, 1-5) following admission, on a median dose of 80 mg of Methylprednisolone or its equivalent of other systemic corticosteroids per day (IQR, 60-107) for a median duration of 5 days (IQR, 4-7) **(Table 1)**.

### Comparison of the cohort with and without corticosteroid

**Table 1** demonstrates the baseline characteristics and outcomes of the study population separated by corticosteroid treatment. Mean patient age was similar in both the cohorts (corticosteroids, 58.67±13.35 years; non-corticosteroid, 57.18±16.81 years, [P = 0.543]). The corticosteroid cohort had more male patients as compared to the non-corticosteroid cohort (86.67% vs. 69.97%, P = 0.011). There was no difference in the distribution of Hispanics race/ethnicity between two cohorts (corticosteroid, 77.97% vs. non-corticosteroid, 71.03%; P = 0.48). Patients among the non-corticosteroids cohort were more obese than the corticosteroid cohort (46.81% vs. 30.51%, P = 0.033). The prevalence of other comorbidities was similar across both cohorts. Patients in the corticosteroid cohort had a low median SpO_2_/FiO_2_ (SF ratio) of 190 (IQR, 92.5-298.44) compared to the median SF ratio of 339.29 (IQR, 278.13-419.05) in the non-corticosteroid cohort (P < 0.001). More percentage of patients in the corticosteroid cohort received Tocilizumab and therapeutic dose of enoxaparin compared to the non-corticosteroid cohort (18.33% vs. 4.83%, P = 0.002; 66.67% vs. 24.83%, P = <0.001, respectively). Among the laboratory parameters, median platelet counts (278.5 vs. 200, P = <0.001), and median AST (55 vs. 44, P = 0.048) were higher in corticosteroid cohort compared to non-corticosteroid cohort. Among inflammatory markers, a median D-Dimer (653.5 vs. 377, P = 0.004) and C-reactive protein (21 vs. 12.1, P = 0.005) were higher in corticosteroid cohort compared to non-corticosteroid cohort.

### Comparison of patients with and without Composite primary outcome

A comparison of demographic, clinical, and laboratory parameters of patients with and without the composite primary outcome is demonstrated in **Table 2**. Compared to patients without the primary outcome, patients with the primary outcome were older (53.78±14.78 vs. 64.52±15.46; OR = 1.62 per 10 years of age; 95% CI, 1.30-2.01; P <0.001), more hypertensive (40.74% vs. 67.16%; OR = 2.98; 95% CI, 1.61-5.5; P=0.001) and had more coronary artery disease (5.19% vs. 20.9%; OR = 4.83; 95% CI, 1.85-12.64; P=0.001). Compared to patients without the primary outcome, patients with the primary outcome were more hypoxic as indicated by lower median calculated PF ratio (256.25, IQR (197.22-280.95) vs. 192.86, IQR (71.72-253.57); OR = 0.93 per 10 unit increase; 95% CI, 0.89-0.96; P <0.001), lower median SF ratio (339.29, IQR (266.67-419.05) vs. 240, IQR (104.44-361.91); OR = 0.96 per 10 unit increase; 95% CI, 0.93-0.98; P=0.001), and more critically ill as indicated by more patient with ≥ 5 Sequential Organ Failure Assessment Score (SOFA) (11.11% vs. 25.37%; OR = 2.72, 95% CI, 1.26-5.87; P = 0.011). Compared to patients without the primary outcome, there was a larger percentage of patients with the primary outcome received a therapeutic dose of Enoxaparin (27.41% vs. 55.22%; OR = 3.27; 95% CI 1.7-6.03; P <0.001). Compared to patients without primary outcome, patients with primary outcome had elevated median value of all inflammatory markers such as D-dimer (389, IQR (238-798) vs. 878.5, IQR (336-3372); OR = 1.05 per 1000 unit increase; 95% CI, 1.001-1.10; P=0.045), and C-reactive protein (11.2, IQR (6.69-22.14) vs. 21.53, IQR (12.95-29.78); OR = 1.06; 95% CI, 1.02-1.10; P=0.003).

**Table 2.** Clinical and Laboratory indices of patients with and without Composite Primary Outcome (ICU transfer or Intubation or Death) **Abbreviations:** BMI – Basal Metabolic Index, CAD – coronary artery disease, HF – Heart failure, Hb – Hemoglobin, WBC – white blood cell, PaO2 – Partial Pressure of Arterial Oxygen, SpO2 = Saturation of oxygen from Pulse oximeter, AST – Aspartate Transferase, ALT – Alanine Transferase, LDH – Lactate Dehydrogenase, IL-6 – Interleukin-6, ICU – Intensive Care Unit, SOFA – Sequential organ failure assessment, IQR – Interquartile range *Per 10 unit Increase, **per 100 unit increase, ***per 1000 unit increase $ - calculated by converting SpO_2_ to PaO_2_, then diving by FiO_2_ # - includes Hydrocortisone, Prednisone, Methylprednisolone, Dexamethasone

|  | Primary Outcome<br>(N=67) | Without Primary<br>Outcome<br>(N=135) | Odds Ratio<br>(95% CI) | P-value |
| --- | --- | --- | --- | --- |
| Demographic Characteristics |  |  |  |  |
| Age, mean (SD) | 64.52 (15.46) | 53.78 (14.78) | 1.62 (1.30-2.01)* | <0.001 |
| Sex, n (%) |  |  |  |  |
| Male | 47 (70.15) | 103 (76.30) | 0.73 (0.38-1.41) | 0.348 |
| Female | 20 (29.85) | 32 (23.70) | Reference |  |
| Ethnicity/Race, n (%) |  |  |  |  |
| Hispanic | 51 (76.12) | 96 (71.64) | 1.29 (0.66-2.54) | 0.452 |
| Comorbidities |  |  |  |  |
| Obesity, n (%) | 26 (39.39) | 57 (43.51) | 0.84 (0.46-1.54) | 0.581 |
| BMI (kg/m<sup>2</sup>), median (IQR) | 28.88 (25-31.39) | 28.64 (24.86-34.83) | 0.99 (0.95-1.04) | 0.740 |
| Hypertension, n (%) | 45 (67.16) | 55 (40.74) | 2.98 (1.61-5.5) | 0.001 |
| Diabetes Mellitus, n (%) | 30 (44.78) | 51 (37.78) | 1.34 (0.74-2.42) | 0.34 |
| COPD/Asthma, n (%) | 14 (20.9) | 18 (13.33) | 1.71 (0.8-3.71) | 0.169 |
| Cancer, n (%) | 5 (7.46) | 4(2.96) | 2.64 (0.69-10.18) | 0.158 |
| CAD, n (%) | 14 (20.9) | 7 (5.19) | 4.83 (1.85-12.64) | 0.001 |
| HF, n (%) | 2 (2.99) | 5 (3.70) | 0.8 (0.15-4.24) | 0.793 |
| Clinical Parameters |  |  |  |  |
| PaO2/FiO2, median<sup>§</sup> (IQR) | 192.86 (71.72-253.57) | 256.25<br>(197.22-280.95) | 0.93 (0.89-0.96)* | <0.001 |
| SpO2/FiO2, median (IQR) | 240 (104.44-361.91) | 339.29<br>(266.67-419.05) | 0.96 (0.93-0.98)* | 0.001 |
| SOFA score < 5, n (%) | 50 (74.63) | 120 (88.89) | Reference | 0.011 |
| SOFA score ≥ 5, n (%) | 17 (25.37) | 15 (11.11) | 2.72 (1.26-5.87) |  |
| Medications |  |  |  |  |
| Corticosteroid<sup>#</sup>, median (IQR) |  |  |  |  |
| Initiation from admission<br>(Days) | 3 (1-4) | 2 (1-5) |  |  |
| Number of patients, n(%) | 13 (19.40) | 45 (33.33) | 0.48 (0.24-0.97) | 0.042 |
| Dose, (mg/day) | 60 (53.5-80) | 80 (60-107) |  |  |
| Duration (Days) | 6 (4-9) | 5 (5-6) |  |  |
| Total Dose (mg) | 320 (192-642) | 400 (240-640) |  |  |
| Hydroxychloroquine, n (%) | 67 (100) | 131 (97.04) | NA | NA |
| Tocilizumab, n (%) | 8 (11.94) | 9 (6.67) | 1.89 (0.70-5.17) | 0.21 |
| Enoxaparin therapeutic dose,<br>n (%) | 37 (55.22) | 37 (27.41) | 3.27 (1.7-6.03) | <0.001 |

| **Laboratory Parameters** |  |  |  |  |
| --- | --- | --- | --- | --- |
| Hb (g/dL), mean (SD) | 12.62 (2.33) | 12.97 (1.96) | 0.92 (0.8-1.06) | 0.269 |
| WBC ($\times 10^3/\mu\text{L}$), median (IQR) | 9.54 (6.16-12.99) | 7.65 (5.8-9.76) | **1.12 (1.04-1.21)** | **0.003** |
| Absolute Lymphocyte Count ($\times 10^3/\mu\text{L}$), median (IQR) | 0.80 (0.58-1.12) | 0.96 (0.69-1.36) | 1.02 (0.78-1.32) | 0.892 |
| Platelets ($\times 10^9/\text{L}$), median (IQR) | 194 (152-260) | 229 (176-323) | **0.96 (0.93-0.99)\*** | **0.011** |
| GFR (ml/min/1.73m<sup>2</sup>), median (IQR) | 60 (34.6-85.5) | 88.6 (60-120.4) | **0.87 (0.81-0.94)\*** | **<0.001** |
| AST (units/L), median (IQR) | 47.5 (37.5-66) | 45 (32-66) | 1.03 (0.96-1.10)* | 0.428 |
| ALT (units/L), median (IQR) | 30.5 (23.5-52) | 34 (21-70) | 0.94 (0.88-1.02)* | 0.173 |
| **Inflammatory Markers** |  |  |  |  |
| Procalcitonin (ng/mL), median (IQR), (N=157) | 0.56 (0.25-1.36) | 0.23 (0.13-0.56) | 0.99 (0.94-1.07) | 0.964 |
| Ferritin (ng/mL), median (IQR), (N=147) | 1037 (821-1685) | 918.5 (427-1690) | 0.98 (0.85-1.12)*** | 0.731 |
| LDH (U/L), median (IQR), (N=145) | 433.5 (347.5-532) | 383 (289-471) | 1.10 (0.94-1.30)** | 0.212 |
| D-Dimer (ng/dL), median (IQR), (N=161) | 878.5 (336-3372) | 389 (238-798) | **1.05 (1.001-1.100)\*\*\*** | **0.045** |
| C-Reactive Protein (mg/dL), median (IQR), (N=132) | 21.53 (12.95-29.78) | 11.2 (6.69-22.14) | **1.06 (1.02-1.1)** | **0.003** |
| IL-6 (pg/ml), median (IQR), (N=35) | 90.5 (32-161) | 85.2 (25.8-131.5) | 1.03 (0.97-1.08)* | 0.385 |

### Primary outcome (composite of ICU transfer, intubation or death)

Out of 202 eligible patients, 13 (22.41%) patients in the corticosteroid cohort developed primary outcome compared to 54 (37.5%) patients developed primary outcome in the non-corticosteroid cohort (P = 0.039). In both unadjusted and adjusted analysis, patients who received corticosteroids were less likely to have had a primary outcome than were patients who did not receive corticosteroids [(unadjusted hazard ratio, 0.45; 95% CI, 0.24 to 0.82; P - 0.009), (**Table 3, Figure 1, Panel A**), (adjusted hazard ratio, 0.15; 95% CI, 0.07 to 0.33; P < 0.001) (**Table 3**)]. Proportionality assumption was not violated (global test P = 0.153).

**Table 3.**
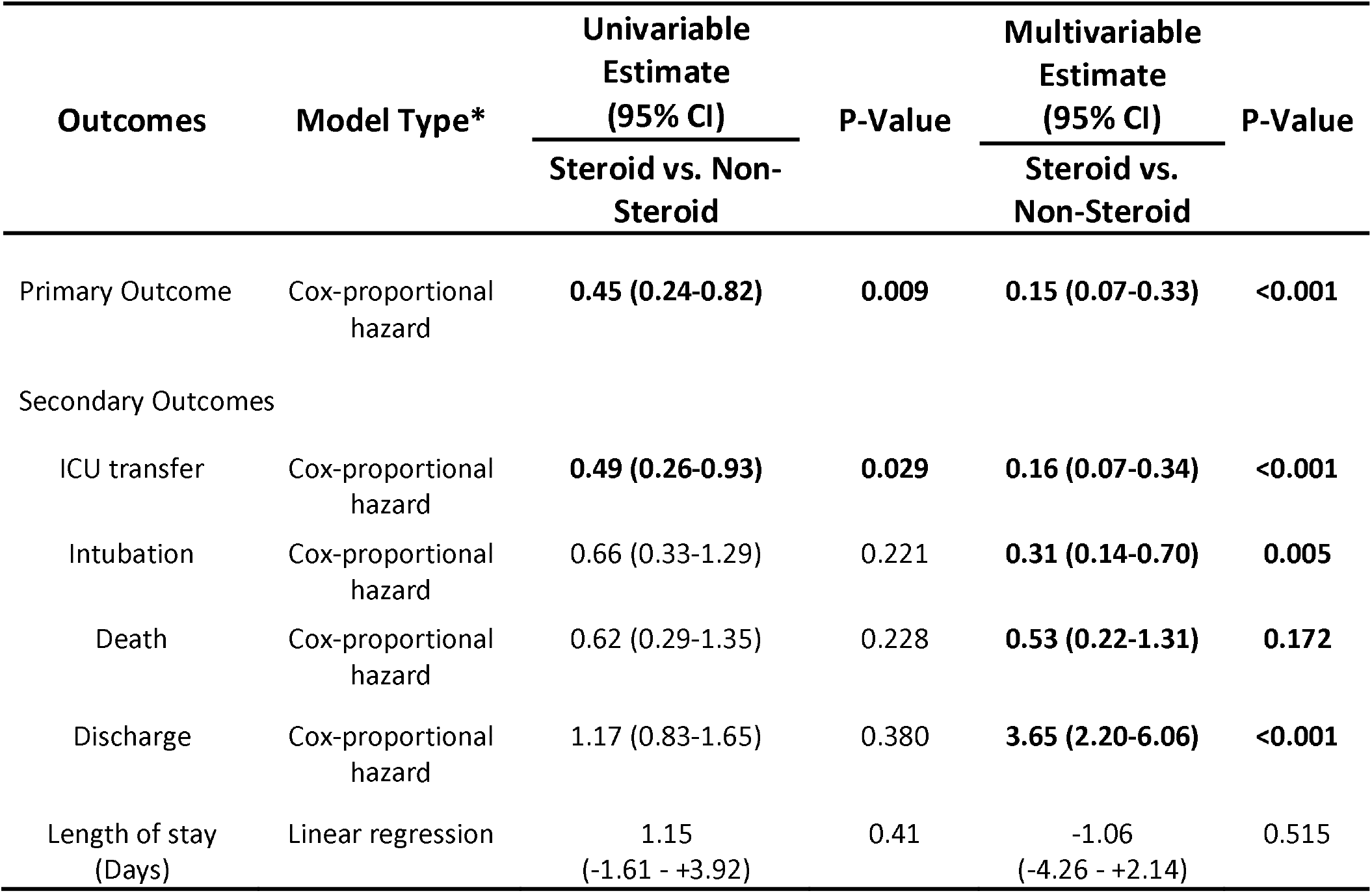
Unadjusted and Model-Adjusted Risk of Primary and Secondary outcome. Primary outcome is composite of ICU transfer, intubation or death *Models adjusted for SF ratio, age, gender, COPD, WBC, platelet count, tocilizumab and therapeutic dose of Enoxaparin

**Figure 1.**
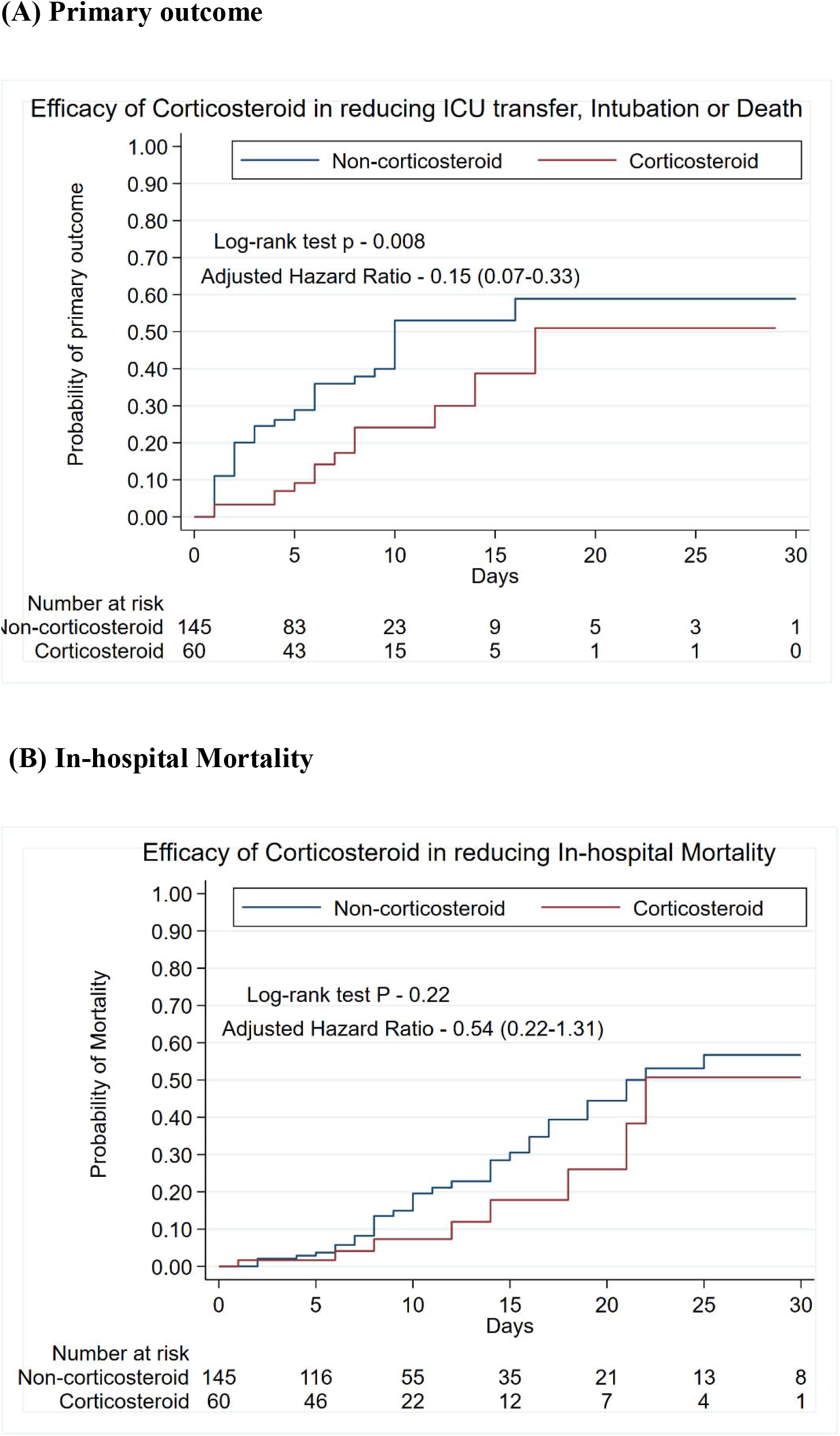
Kaplan-Meier curve of patients with non-severe COVID-19 pneumonia who received and did not receive corticosteroids. **Panel (A)** Primary Outcome; **Panel (B)** In-hospital Mortality Primary outcome is a composite of ICU transfer, intubation or death

### Secondary outcomes

In the corticosteroid cohort, 12 (20.69%) patients were transferred to ICU as compared to 47 (32.64%) patients in the non-corticosteroid cohort (P = 0.09). In the unadjusted and adjusted Cox-analysis, patients who received corticosteroid were less likely to require ICU transfer than were patients who did not receive corticosteroids [(unadjusted hazard ratio, 0.49; 95% CI, 0.26 to 0.93; P – 0.029), (adjusted hazard ratio, 0.16; 95% CI, 0.07 to 0.34; P < 0.001)] (**Table 3, Supplemental Appendix 4**). In the corticosteroid cohort, 11 (18.97%) patients were intubated compared to 36 (25.35%) patients in the non-corticosteroid cohort (P = 0.33). In the unadjusted and adjusted Cox-analysis, patients who received corticosteroid were less likely to require intubation than were patients who did not receive corticosteroids [(unadjusted hazard ratio, 0.66; 95% CI, 0.33 to 1.29; P – 0.221), (adjusted hazard ratio, 0.31; 95% CI, 0.14 to 0.70; P – 0.005)] (**Table 3, Supplemental Appendix 5)**. In the corticosteroid cohort, 8 (14.55%) patients died compared to 34 (25%) patients in the non-corticosteroid cohort (P = 0.09). In the unadjusted and adjusted Cox-analysis, patients who received corticosteroid were less likely to die than were patients who did not receive corticosteroids [(unadjusted hazard ratio, 0.62; 95% CI, 0.29 to 1.35; P – 0.228), (adjusted hazard ratio, 0.53; 95% CI, 0.22 to 1.31; P – 0.172)]; however, it was statistically non-significant (**Table 3, Figure 1, Panel B)**. In the corticosteroid cohort, 47 (85.45%) patients were discharged compared to 102 (75%) patients in non-corticosteroid cohort (P = 0.11). In the unadjusted and adjusted Cox-analysis, patients who had received corticosteroid had more probability of being discharged than were patients who did not [(unadjusted hazard ratio, 1.17; 95% CI, 0.83 to 1.65; P – 0.380), (adjusted hazard ratio, 3.65; 95% CI, 2.20 to 6.06; P < 0.001)] (**Table 3, Supplemental Appendix 5**). Proportionality assumptions were not violated for any of the secondary outcomes tested by Cox-regression. The median length of stay was higher in the corticosteroid cohort (9 days, IQR (6-17)) compared to the non-corticosteroid cohort (7 days, IQR (5-13.5); P = 0.025) **(Table 1, Table 3)**. Using multivariable linear regression, length of stay was lower in corticosteroid cohort but statistically non-significant (coefficient −1.06; 95% CI, −4.26 to 2.14; P=0.515).

### Daily Trend of SpO_2_/FiO_2_ since the Index date

**Figure 2** demonstrates the comparison of a daily trend of SF ratio between patients with and without corticosteroids. The graph depicts that the corticosteroid cohort had lower baseline SF ratios (b=-185.97, p<0.001), SF ratio was found to increase over time (b=24.48, p=0.025). Furthermore, significant interactions between treatment and time demonstrated that the non-corticosteroid cohort experienced a decrease in SF ratio over time compared to the corticosteroid cohort, who were found to experience an increase in SF ratio over time. The likelihood ratio test vs. linear model chi-square test indicated that the use of a multilevel model was appropriate for the data.

**Figure 2.**
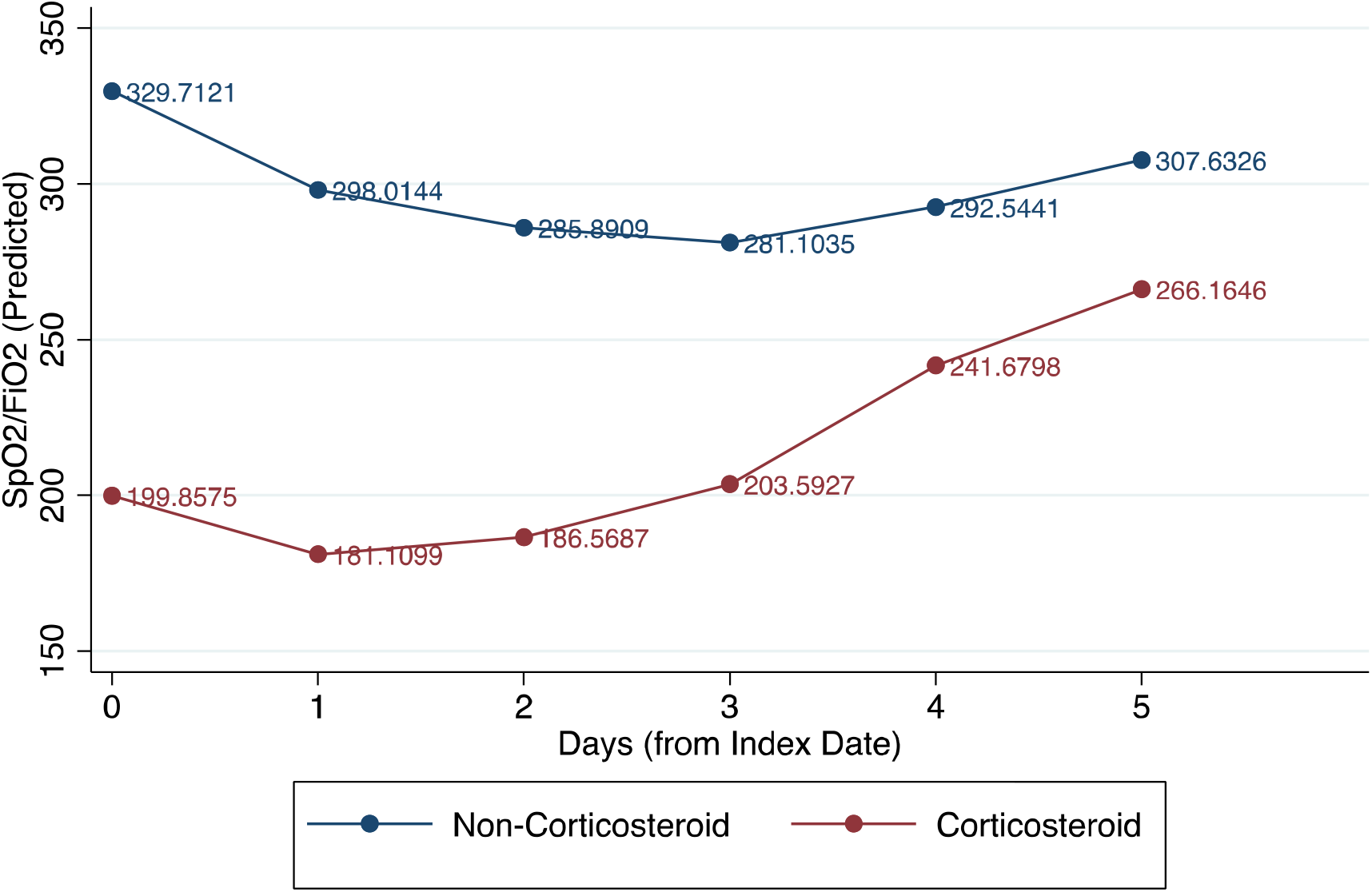
Comparison of trend of mean SpO2/FiO2 Ratio since Index date with and without corticosteroid.

## Discussion

The results of our study showed that the administration of corticosteroids in patients admitted to the general medical ward with AHRF and a diagnosis of COVID-19 pneumonia was associated with a lower risk of developing the primary outcome of ICU transfer, intubation or death. Regarding secondary outcomes, patients who received corticosteroids were found to have a lower risk of ICU transfer, intubation, in-hospital death, and were more likely to be discharged. In the corticosteroid cohort, however, a lower hazard of mortality did not show statistical significance likely due to smaller sample size.

As we noted in the introduction, the findings from an earlier study have shown the controversial role of corticosteroids in COVID-19. A more recent study by Wu et al. demonstrated a possible beneficial role of methylprednisolone in reducing the risk of death in the subgroup analysis of 84 patients with ARDS in an observational study of 201 patients with COVID-19.^13^ A meta-analysis of randomized clinical trials suggested corticosteroid could reduce mortality and need for mechanical ventilation in patients with severe community-acquired pneumonia, irrespective of bacterial or viral etiology.^14,15^ A preliminary report from the RECOVERY trial showed a beneficial role of low-dose of dexamethasone in reducing mortality among patients on mechanical ventilation and requiring oxygen therapy but no benefit among patients who did not require oxygen therapy in patients with COVID-19 (NCT04381936). On the contrary, a meta-analysis showed higher mortality among patients receiving corticosteroids.^16^ However, this meta-analysis included patients with SARS, MERS, COVID-19, and had high heterogeneity. A subgroup analysis from the same study reported no significant difference in mortality between two cohorts when included only COVID-19 patients ^16^, suggesting corticosteroids may not be harmful in these patients. We assume studies that showed higher or no difference in mortality with corticosteroid were critically ill with ARDS, on mechanical ventilation, and they might have passed the point where adverse outcomes could be modified by corticosteroid.

In COVID-19 pneumonia, lung injury is associated with a direct virus-induced injury. However, the severity of illness is associated with the virus triggered immune hyper-response that is characterized by activation of various immune cells and release of numerous pro- and anti-inflammatory cytokines, including tumor necrosis factor (TNF), Interleukin-6 (IL-6), and many more. Overwhelming secretion of cytokines leads to severe lung damage manifested as the destruction of the small airway, alveolar epithelium, and vascular endothelium that progresses to pulmonary edema and hyaline membrane formation.^17,18^ In severe COVID-19 pneumonia, patients’ symptoms worsen and become more hypoxic during the 4-7 days after onset of symptoms. ^19^ Hence, it is vital to suppress the cytokine storm before that period. We, therefore, believe a majority of patients survive and recover if they overcome the period of the cytokine storm. Corticosteroid is the classical immunosuppressive drug that helps in delaying or halting the progress of pneumonia and has been effective for the treatment of ARDS. ^20,21^ In addition to immunosuppressive properties, corticosteroid possesses anti-inflammatory activity that reduces systemic inflammation, decreases exudation into the lung tissue, promotes the absorption of inflammation, and prevents alveolar damage. ^22^ These effects of corticosteroid help in relieving hypoxemia earlier, preventing further progression of respiratory insufficiency, and hence associated with improved primary, secondary outcomes, and SF ratio in the study.

This study was also unique in that the average duration of usage of corticosteroids was 5 days, and the treatment population was not intubated or admitted to an ICU. Traditionally corticosteroids are not used early in viral pneumonia due to concerns of delayed viral clearance. However, delayed treatment may predispose to worsening and progressive inflammatory response and multiorgan failure. Hence, the authors presume that careful monitoring of inflammatory markers might help us to guide the judicious use of corticosteroid. This study highlights that early administration of corticosteroid in COVID-19 viral pneumonia may not be as harmful as initially suspected. On the contrary, the authors found a significantly beneficial role of corticosteroid in lowering transfer to ICU, risk of intubation, and in-hospital mortality. The ICU transfer and Intubation might be linked to the quality of life of the patient. Since the study showed a lower risk of ICU transfer, intubation risk, and a higher probability of discharge among patients treated with corticosteroid, corticosteroid might be associated with improved quality of life in the early period after receiving corticosteroid. However, data on readmission to hospital or ED visits post-discharge would be required to check this presumptive role.

## Limitations

This study has several limitations, given the observational nature of the study. It is a single-center study, and most of the patients were Hispanic that limits the generalizability of the study. There were missing data for Inflammatory markers and potential inaccuracies in the documentation of variables in the electronic health records; however, as mentioned above, two authors made sure the accuracy of the collected database. Due to missing data for inflammatory markers, adjustment with those variables was not possible. Despite the extensive adjustments, it is still possible that unmeasured confounding prevails. We did not have longitudinal data with follow-up. A randomized clinical trial is the best approach to determine whether the benefit can be ascribed to any given therapeutic intervention as trial design diminishes the two major hurdles of observational studies, namely unmeasured confounding and bias. One randomized clinical trial from China has been registered in which patients were randomly assigned to receive 1 mg/kg methylprednisolone (NCT04273321). The authors tried to minimize confounding by choosing the best possible model in multivariable regression in this retrospective cohort study.

## Conclusions

In our analysis of hospitalized patients in the general ward with COVID-19 pneumonia complicated by acute hypoxic respiratory failure, earlier use of low dose systemic corticosteroid was associated with a significantly lower risk of the primary outcome of ICU transfer, intubation, or in-hospital death. Given the observational design, the study should be interpreted with caution due to potential bias and residual confounders. Double-blinded randomized clinical trials should be conducted to validate these results.

## Data Availability

Not available given the identifiable nature of the dataset

## Acknowledgment

We acknowledge the dedication, commitment, and sacrifice of the staff, providers, and personnel in our institution through the Covid-19 crisis and the suffering and loss of our patients as well as in their families and our community. We acknowledge Ms. Ejiro C. Gbaje MPH for her help in the analysis of the data.

## Abbreviation

COVID-19: Coronavirus disease 2019
AHRF: Acute Hypoxemic Respiratory Failure
ICU: Intensive Care Unit
SF ratio: SpO_2_/FiO_2_ ratio

